# A robust continuous wavelet transform (CWT) based for R-peak detection method of ECG

**DOI:** 10.1101/2023.07.31.23293050

**Authors:** Lola El Sahmarany, Maha Alshammari, Mahbubunnabi Tamal, Abdulhakeem Alomari

## Abstract

Cardiovascular disease is the main cause of death worldwide. An electrocardiogram (ECG) signals is typically used as the first diagnosis tool to detect abnormality in the heart signal. Reliable detection of R-peak in the ECG signal indicates various heart malfunctions (e.g., arrhythmia) and allows for proactive prevention of death due to cardiovascular disease. Though several R-peak detection methods have been proposed, the existence of noise in ECG signals and changes in QRS morphology compromise the robustness and reliability of these methods. Therefore, the need for a reliable detection of R-peak is crucial for diagnosis of heart abnormalities. This paper introduces a time-efficient and novel continuous wavelet transform (CWT) based method for R-peak detection. The proposed method first transforms the ECG signal in to time-frequency spectrum. The contributions of different frequencies at every time point are then calculated from the time-frequency spectrum to efficiently reduce the impact of noise and generate a summed frequency signal. A threshold technique is also proposed to detect the R-peak from the newly generated signal allows. The MIT-BIH arrhythmia database is used as a reference for validation and comparison of the proposed method with the results of other existing R-peak detection methods. The experimental results prove the efficiency and robustness of the developed method on noisy ECG signals with changes in QRS morphology with 99.87% sensitivity, 99.76% positive prediction value and a detection error rate of only 0.37%. In addition to the high accuracy in detecting R-peaks, the ease-to-use and fast-processing make the proposed method an efficient and reliable tool for real-time abnormality detection in ECG signal.

## 1. Introduction

Human body has a considerable number of physiological parameters that represent the conditions of its various organs. Worldwide, the mortality rate from cardiovascular disease is very high [1]. The main parameter to diagnose cardiovascular anomalies is the heart rate that measures the number of heart beats per minute. Electrocardiogram (ECG) signals is the most commonly among various physiological signals for extracting the heart rate. ECG signal is obtained invasively by recording the electrical potential directly on the heart or using electrodes. Medical doctors use the electrocardiogram as graphical temporal representation which contains considerable amount of information on functioning and possible pathologies of heart [2].

ECG signal represents the electrical activity of heart which is an essential indicator in the diagnosis of cardiovascular diseases [2]. One of the main elements of ECG is QRS complex that represents the largest wave in ECG signal. Width and shape of QRS complexes collectively provide relevant information on condition of heart and its ventricular function [3].

However, various signals can interfere with the measurement of an ECG such as electrical EMG signals circulated in nerves, artefacts due to movement of person or displacement of electrodes. Such interference may affect the accuracy of ECG signal measurement due to the relatively small amplitude of ECG waves (i. e. few millivolts) which may also vary from a person to another [4]. Although R-peak has the highest amplitude of ECG signal, however, it only ranges between 0.5mV and 4mV if it is measured near the heart. Thus, the measurement of R-peak amplitude is questionable especially when considerable amount of noise is involved.

In QRS segment, R-peak is characterized by short duration with sharp transition compared to other elements of ECG Signal. Normally, R-peak presents a sinus rhythm, and it is easily detectable by its amplitude in an ECG recording of good quality. However, this is not the case for atypical ECG signals where the R-peak sometimes presents a low amplitude. Literature highlights various technique for R peak detection as follows: Digital filtering methods [5], [6], Shannon energy envelope (SEE) based methods [7], [8], Slope-based threshold methods [9], Wavelet-transform-based methods [10]– [11], and Mathematical morphology-based methods [10], [12].

The aforementioned techniques show more or less satisfactory results depending on the context. However, the implementation of an effective QRS detector is considerably challenging regardless of the type of ECG recording or the nature of noise involved.

Thus, the commonly used techniques for QRS detection in literature will be discussed to highlight the advantages and disadvantages of each relevant techniques.

The Pan and Tompkins methods is one of the most common benchmarks for R-peak detection that incorporates five basic techniques including; low-pass filter and high-pass filter, derivative filter technique, squaring, and windowing for the detection of the R peaks [13][14]. Using high quality ECG signals, these methods could obtain high detection accuracy, however, the accuracy of the Pan-Tompkins based QRS detection methods is questionable on a low-quality ECG database.

Wavelet transforms methods (WT) are also developed for detecting QRS complexes. These methods are based on multiscale features used to distinguish QRS complexes from P-T waves or noise.

Martinez et al. [15] proposed a global wavelet-based approach to detect the QRS complex. Then, the detected QRS is delineated to determine the start and end of the QRS complex peak. For detection of the QRS complex, the authors integrate the multiscale maximum modulus approach with thresholding to determine the local maxima in ECG signal.

Also, in [16] authors choose to combine the derivatives Hilbert transform, wavelet transform, and adaptive thresholding to enhance the QRS complex with respect to other waves and artifacts in ECG signal. In [8], another combination of methods, Shannon energy (SE) method with Hilbert transform method [17] provide good results for detecting QRS complex. However, the above combination methods are unsuitable for real-time application due to Hilbert transform methods which requires large processing time and memory. Moreover, these methods detect several noise peaks in ECG signal with long pauses.

It is noted from the literature that, most of QRS detection methods are developed based on three steps denoising of the ECG signal, QRS complex enhancement and decision rule for peaks detection. Some researchers [18] utilized time domain techniques for processing ECG signals however, these methods yield to low accuracy when noise or artifact is involved. Other researchers [19] developed frequency domain techniques that are not suitable for real time pathological datasets due to spectral leakage problems which lead to a high rate of false positive detection. In addition, frequency and time domain-based techniques are not effective in analyzing ECG signals with non-linear behavior. For that, most of the pre-process methods need to be designed carefully, and several nonlinear transform techniques are necessary to enhance the QRS complex.

According to the above literature, there is a need for developing a new method to analyze ECG signals with nonlinear and non-stationary features. This paper introduces a novel R-peak detection method that integrates continuous wavelet transform (CWT) with the summation of frequency contribution at each time point of CWT and moving average to rapidly detect the R peaks in QRS complex.

In the remaining part of this paper, a new method will be proposed to improve the detection performance of R-peaks. The proposed method will be applied to real case studies to evaluate its performance. Also, the implementation results will be presented, analyzed, and discussed. Finally, conclusion will be drawn along with recommendation for future work.

## 2. Materials and Methods

### 2.1. Proposed R-peak detection method

The block diagram in Figure 1 presents the proposed method for detecting the R-peak. In this method first, a time-frequency spectrum is generated using continuous wavelet transforms (CWT). Wavelet Transform (WT) decomposes the ECG signal in the time-frequency components to characterize the signal at different resolutions and is considered as the most efficient tool for solving noise problem [11]. The main advantage of wavelets is to work in sub-band which allows the separation of noise components from useful signal components.

**Figure 1.**
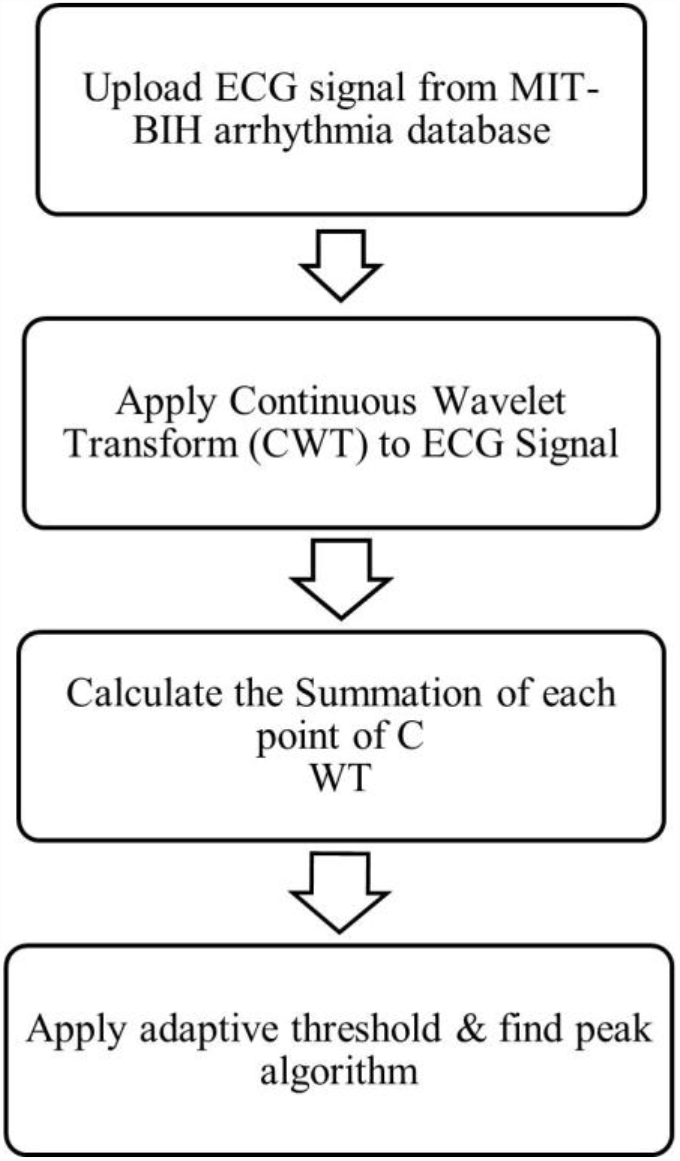
Flowchart for R-peak detection processes

The contributions of different frequencies at every time point are then calculated from the time-frequency spectrum to efficiently reduce the noise and generate a summed frequency signal. The detail of the proposed method is provided below.

If *f*(*t*) represents the obtained original signal, the continuous wavelet transforms CWT of *f*(*t*) at a scale factor (dilation) related to the notion of frequency (*a* > 0) and offset (translation), related to the notion of position temporal *b* can be expressed as [10]:

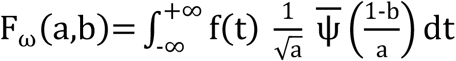

where *ψ* (*t*) *is* the continuous wavelet function called the mother wavelet, 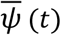 is the complex conjugate of and 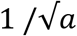 the normalization factor.

Then, summation of *F*_*ω*_ (*a, b*) at every time point *b* that represents the contribution of all frequencies at each *b* is expressed as:

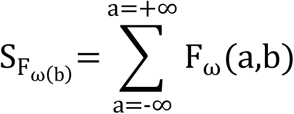

The peak detection phase attempts to accommodate the variation and morphology of the amplitude changes of ECG signals complexes using a threshold-based algorithm. To identify R-peak signal, a sliding window of 1000 samples is applied to 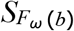. Then, the mean value of the maximum of R-peak in each window is calculated in order to determine the threshold values. The use of this technique is to avoid that the threshold being dominated by a large or small extreme value in 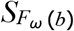. Because of multiple candidate R-peaks can be present within the same QRS complex and the candidate QRS peaks also include sharp P and T-peaks, we propose the following decision rules to identify the most prominent R-peaks. The threshold is expressed as follow:

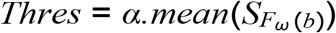

Where *α* is denotes the constant range between [0.9-1.5] that multiplies the mean value of the samples of each window.

Figure 2 shows an example of the simulation result of time-frequency spectrum *F*_*ω*_(*a, b*)of the ECG signal (Figure 2. i) and the summation of frequency contribution at each time point *F*_*ω*_(*b*) (red) as shown in Figure 2 (ii). The locations of positive peaks which are referred to the locations of true R-peaks are clearly identified in Figure 2 (iii).

**Figure 2.**
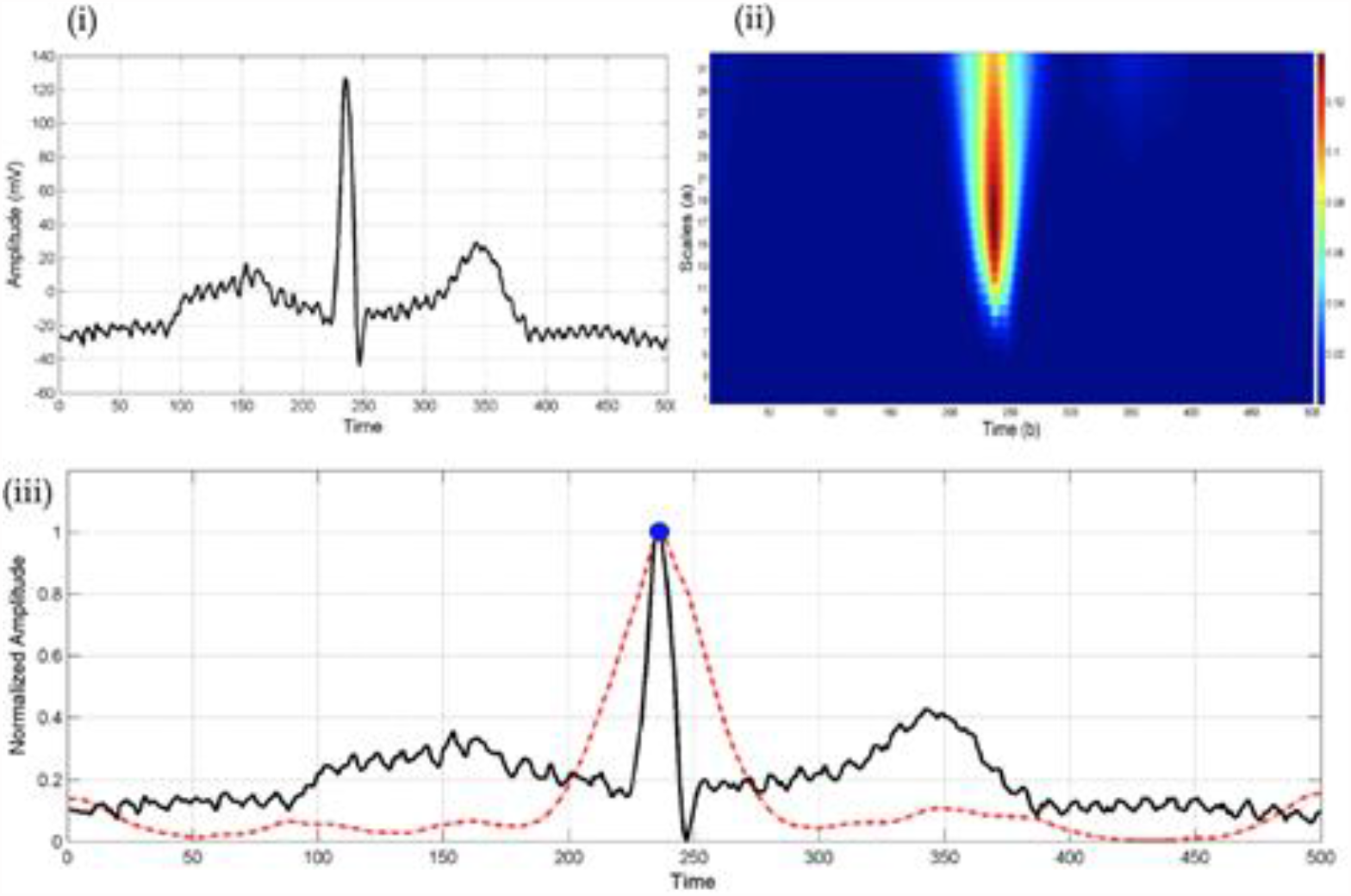
Detection of R-peak from ECG signal. (i) Original ECG signal *f*(*t*). (ii) Time-frequency spectrum *F*_*ω*_ (*a, b*) of the ECG signal. (iii) ECG signal (black) along with the summation of frequency contribution at each time point *F*_*ω*_ (*b*) (a) (red)

Figure 3 shows the total signal processing of the proposed CWT method for detecting R-peaks applied on record #121. After the summation of frequency contribution of each point time of CWT, the threshold operation method is applied to detect the R-peak. In Figure 3. iv, the locations of positive peaks are referred to as the locations of true R-peaks.

**Figure 3.**
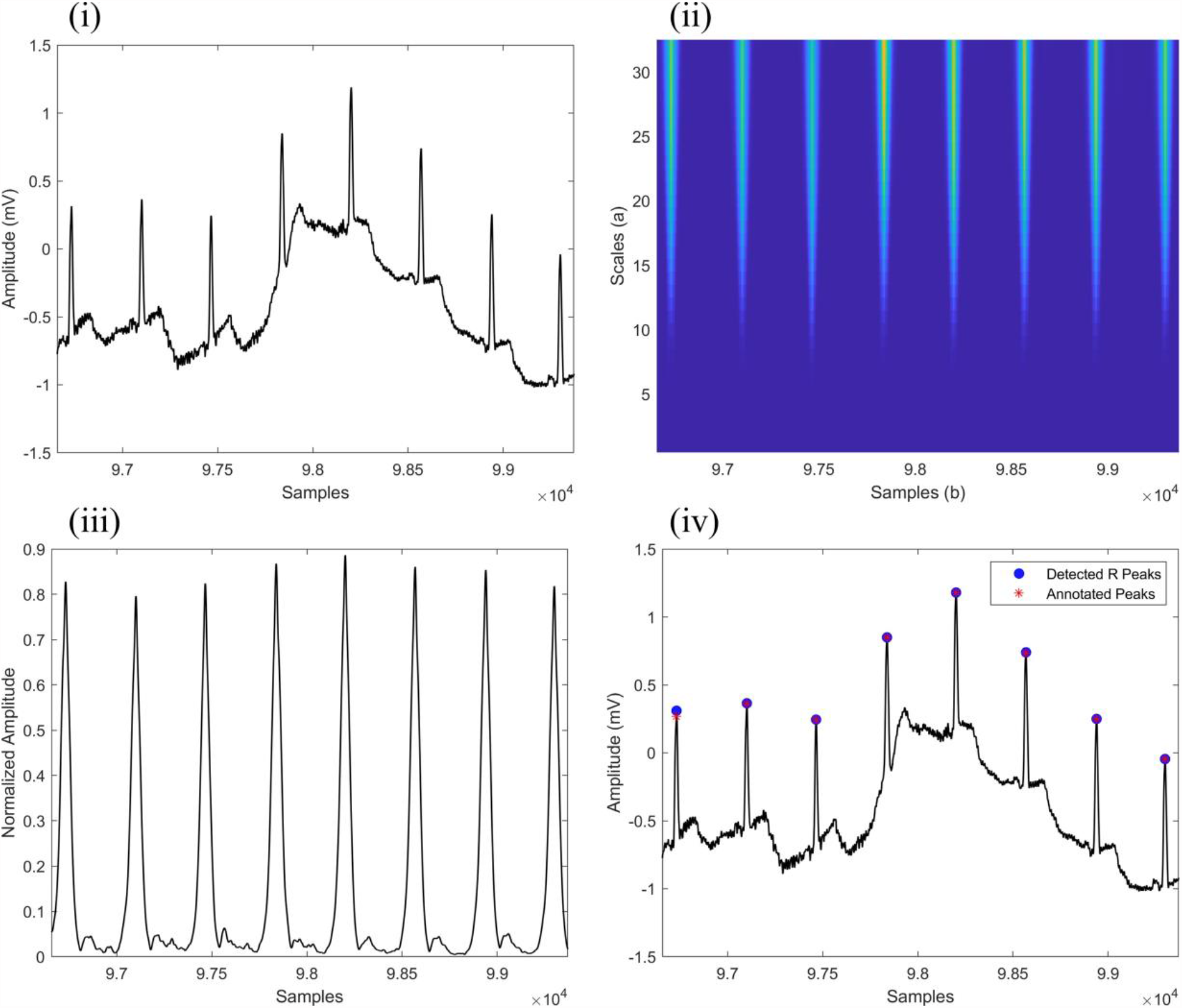
Example of the proposed R peak detection method using CWT and its summation of frequency contribution of each point time. (i) Input ECG signal of record 121, (ii) Time-frequency spectrum *F*_*ω*_(*a, b*) of the ECG signal, (iii) result of frequency contribution of each point time, (iv) true detected R peaks in ECG signal (blue circles) and annoted Peaks by the expert (red stars).

### 2.2. Performance evaluation of the proposed method

The main objective of this paper is to develop a clinical support tool for the medical profession that allows for accurate detection of R-peak of QRS waves. The proposed method was implemented using Matlab R2020b^®^ on a laptop with a 2.3 GHz 8-Core Intel Core i9, 16 GB of memory on macOS on a real ECG signal MIT-BIH database [20]. The MIT-BIH Arrhythmia is the most widely used database that contains 48 half-hour recordings of ECG signals which are manually annotated by experts (sampling rate: 360Hz, resolution: 11 bits over a range of 10 mV). The database recordings correspond to 25 men aged between 32 and 89, and 22 women aged between 23 and 89. However, some of these ECGs (e.g. #100 and #107) contain well identified R peaks and other very interesting physiological information, others contain difficult to detect QRS complexes and abnormal shapes, noise and artifacts (e.g. #105, #108, #116, #203 and #207). Recordings of MIT-BIH Arrhythmia database represent a reference for validation and comparison of detection algorithms on ECG signals. The main advantage of MIT-BIH database is the inclusion of a large number of cardiac pathologies that allows for validation of detection algorithms on a large number of cases.

It should be noted that each recording has been annotated by experts and cardiologists to increase the reliability of data. If an identified QRS peak equals to annotated time, ± 100 *ms* then it will be detected as a true peak [21] [22].

## 3. Results and Discussion

The algorithm’s ability for QRS detection is evaluated using binary classification. This classification is based on the three selection states allowing to describe the detected and /or undetected peaks as follows:

- True Positive (TP) is the number of correct QRS peaks detected as true peaks.
- False Positive (FP) is the number of incorrect QRS peaks detected as true peaks.
- False Negative (FN) is the number of incorrect QRS peaks undetected as true peaks.

The selection states allow for evaluating the efficiency of algorithm in detecting the QRS peaks using three indices: sensitivity (Se), positive prediction value (+P) and detection error rate (DER). These indices can be calculated using the following equations in Table 1.

**Table 1:**
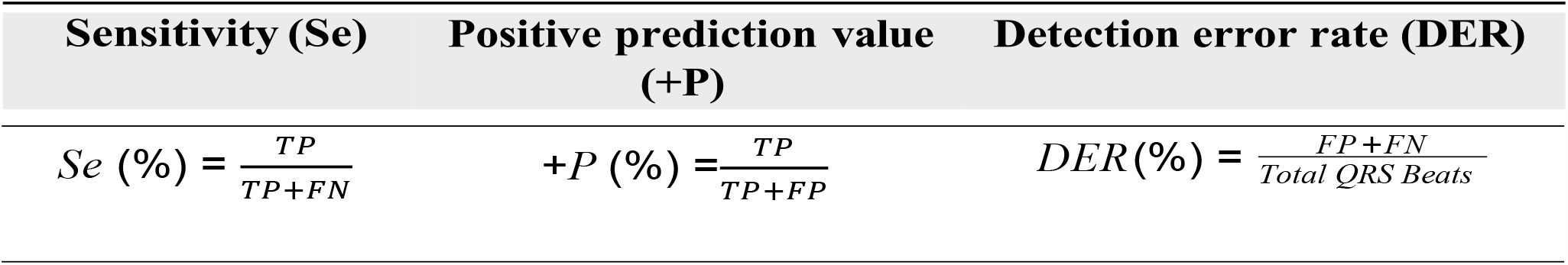
Three metrics used to evaluate the performance of R-peak detection were.

| Sensitivity (Se) | Positive prediction value (+P) | Detection error rate (DER) |
| --- | --- | --- |
| $Se (\%) = \frac{TP}{TP+FN}$ | $+P (\%) = \frac{TP}{TP+FP}$ | $DER(\%) = \frac{FP+FN}{Total\ QRS\ Beats}$ |

Table 2 summarizes the performance of the proposed method using the MIT-BIH arrhythmia database. Also, it illustrates the ability of the proposed method to detect the most of QRS peaks regardless of the existence of abrupt changes, baseline drift, low amplitude QRS complexes, high-frequency noise, sharp P and T waves, or wide ventricular premature contractions in the ECG signal. The illustrated results show that a total of 409 detection failures includes 141 false-negative beats and 268 false-positive beats. It should be noted that recordings 105, 203, and 207 have the highest number of detection failures with a total of 48 failures, 54 failures, and 61 failures respectively. Note that in record 207 there are 2 min ventricular flutter segments. Therefore, most state-of-the-art QRS identification algorithms [14] [23] [17] [24] eliminate record of 207 or the 2-min ventricular flutter segments due to its strange morphology.

**Table 2:** Summary performance analysis of the proposed method using the MIT-BIH arrhythmia database.

| No. | Time<br>Consumption (s) | Total<br>Beats | TP | FN | FP | FP+FN | Se (%) | +P(%) | DER(%) |
| --- | --- | --- | --- | --- | --- | --- | --- | --- | --- |
| 100 | 1.166 | 2273 | 2273 | 0 | 0 | 0 | 100.00 | 100.00 | 0.00 |
| 101 | 1.1182 | 1865 | 1862 | 3 | 4 | 7 | 99.84 | 99.79 | 0.38 |
| 102 | 1.088 | 2187 | 2187 | 0 | 0 | 0 | 100.00 | 100.00 | 0.00 |
| 103 | 1.091 | 2084 | 2084 | 0 | 0 | 0 | 100.00 | 100.00 | 0.00 |
| 104 | 1.203 | 2229 | 2225 | 4 | 7 | 11 | 99.82 | 99.69 | 0.49 |
| 105 | 1.122 | 2572 | 2552 | 20 | 28 | 48 | 100.00 | 98.91 | 1.87 |
| 106 | 1.158 | 2027 | 2027 | 0 | 2 | 2 | 99.91 | 99.90 | 0.10 |
| 107 | 1.153 | 2137 | 2135 | 2 | 0 | 2 | 99.91 | 100.00 | 0.09 |
| 108 | 1.185 | 1763 | 1756 | 7 | 15 | 22 | 99.60 | 99.15 | 1.25 |
| 109 | 1.128 | 2532 | 2532 | 0 | 0 | 0 | 100.00 | 100.00 | 0.00 |
| 111 | 1.156 | 2124 | 2123 | 1 | 1 | 2 | 99.95 | 99.95 | 0.09 |
| 112 | 1.158 | 2539 | 2539 | 0 | 0 | 0 | 100.00 | 100.00 | 0.00 |
| 113 | 1.158 | 1795 | 1795 | 0 | 13 | 13 | 100.00 | 99.28 | 0.72 |
| 114 | 1.179 | 1879 | 1879 | 0 | 16 | 16 | 100.00 | 99.16 | 0.85 |
| 115 | 1.124 | 1953 | 1953 | 0 | 0 | 0 | 100.00 | 100.00 | 0.00 |
| 116 | 1.098 | 2412 | 2401 | 11 | 7 | 18 | 99.54 | 99.71 | 0.75 |
| 117 | 1.097 | 1535 | 1535 | 0 | 13 | 13 | 100.00 | 99.16 | 0.85 |
| 118 | 1.111 | 2278 | 2278 | 0 | 0 | 0 | 100.00 | 100.00 | 0.00 |
| 119 | 1.173 | 1987 | 1987 | 0 | 0 | 0 | 100.00 | 100.00 | 0.00 |
| 121 | 1.146 | 1863 | 1862 | 1 | 1 | 2 | 99.95 | 99.95 | 0.11 |
| 122 | 1.144 | 2476 | 2476 | 0 | 0 | 0 | 100.00 | 100.00 | 0.00 |
| 123 | 1.131 | 1518 | 1518 | 0 | 0 | 0 | 100.00 | 100.00 | 0.00 |
| 124 | 1.077 | 1619 | 1619 | 0 | 1 | 1 | 100.00 | 99.94 | 0.06 |
| 200 | 1.065 | 2601 | 2598 | 3 | 2 | 5 | 99.88 | 99.92 | 0.19 |
| 201 | 1.093 | 1963 | 1959 | 4 | 2 | 6 | 99.80 | 99.90 | 0.31 |

| *Table 2 continued* |  |  |  |  |  |  |  |  |  |
| --- | --- | --- | --- | --- | --- | --- | --- | --- | --- |
| **No.** | **Time**<br>**Consumption (s)** | **Total**<br>**Beats** | **TP** | **FN** | **FP** | **FP+FN** | **Se (%)** | **+P(%)** | **DER(%)** |
| 202 | 1.149 | 2136 | 2126 | 10 | 1 | 11 | 99.53 | 99.95 | 0.51 |
| 203 | 1.111 | 2980 | 2953 | 27 | 27 | 54 | 99.09 | 99.09 | 1.81 |
| 205 | 1.135 | 2656 | 2629 | 2 | 0 | 2 | 99.92 | 100.00 | 0.08 |
| 207 | 1.123 | 1859 | 1853 | 6 | 55 | 61 | 99.68 | 97.12 | 3.28 |
| 208 | 1.094 | 2954 | 2939 | 15 | 6 | 21 | 99.49 | 99.80 | 0.71 |
| 209 | 1.123 | 3005 | 3005 | 0 | 1 | 1 | 100.00 | 99.97 | 0.03 |
| 210 | 1.110 | 2649 | 2573 | 4 | 3 | 7 | 99.84 | 99.88 | 0.26 |
| 212 | 1.152 | 2748 | 2748 | 0 | 0 | 0 | 100.00 | 100.00 | 0.00 |
| 213 | 1.070 | 3251 | 3245 | 6 | 0 | 6 | 99.82 | 100.00 | 0.18 |
| 214 | 1.147 | 2261 | 2260 | 1 | 19 | 20 | 99.96 | 99.17 | 0.88 |
| 215 | 1.086 | 3363 | 3318 | 0 | 0 | 0 | 100.00 | 100.00 | 0.00 |
| 217 | 1.059 | 2208 | 2206 | 2 | 4 | 6 | 99.91 | 99.82 | 0.27 |
| 219 | 1.150 | 2154 | 2154 | 0 | 0 | 0 | 100.00 | 100.00 | 0.00 |
| 220 | 1.054 | 2047 | 2044 | 3 | 0 | 3 | 99.85 | 100.00 | 0.15 |
| 221 | 1.135 | 2427 | 2426 | 1 | 0 | 1 | 99.96 | 100.00 | 0.04 |
| 222 | 1.145 | 2483 | 2429 | 6 | 11 | 17 | 99.75 | 99.55 | 0.68 |
| 223 | 1.157 | 2605 | 2605 | 0 | 0 | 0 | 100.00 | 100.00 | 0.00 |
| 228 | 1.110 | 2053 | 2052 | 0 | 29 | 29 | 100.00 | 98.61 | 1.41 |
| 230 | 1.130 | 2256 | 2256 | 0 | 0 | 0 | 100.00 | 100.00 | 0.00 |
| 231 | 1.148 | 1571 | 1570 | 1 | 0 | 1 | 99.94 | 100.00 | 0.06 |
| 232 | 1.190 | 1780 | 1779 | 1 | 0 | 1 | 99.94 | 100.00 | 0.06 |
| 233 | 1.150 | 3078 | 3078 | 0 | 0 | 0 | 100.00 | 100.00 | 0.00 |
| 234 | 1.201 | 2753 | 2753 | 0 | 0 | 0 | 100.00 | 100.00 | 0.00 |
| **Total** | **54.251** | **109488** | **109156** | **141** | **268** | **409** | **99.87** | **99.76** | **0.37** |

Some indicators including positive prediction value, sensitivity, and detection error rate have been evaluated to check the detection accuracy of the proposed method.

The results show that the prediction value ranges from 97.12% to 100%, the sensitivity ranges from 98.84% to 100%, and the detection error rate ranges from 0% to 1.87%. Also, the overall positive predictive value, sensitivity, and detection error rate have been evaluated as a total of 99.76%, 99.87%, and 0.37% respectively.

A 30-minute ECG record takes about 1.3 seconds to be processed. The proposed technique can be implemented for wearable heart rate monitoring and automatic ECG analysis, according to the findings.

A comparison between the results of the proposed method and that of existing wavelet-based methods using MIT-BIH arrhythmia database is illustrated in Table 3. The comparison of the proposed method with WT-based methods indicates its comparable performance as illustrated in Table 3.

**Table 3:** Performance comparison of the proposed method with the existing wavelet-based methods for detecting R-peaks using MIT-BIH database.

| Methods | TP | FN | FP | FN+FP | Se (%) | +P (%) | DER (%) |
| --- | --- | --- | --- | --- | --- | --- | --- |
| Proposed method | 109,156 | 141 | 268 | 409 | 99.87 | 99.76 | 0.37 |
| Ghaffari et al., 2007 [10] | 109,837 | 120 | 322 | 440 | 99.91 | 99.72 | 0.40 |
| Chun-Cheng Lin et al. 2019 [22] | 109,308 | 180 | 91 | 271 | 99.84 | 99.92 | 0.25 |
| Zidelmal et al., 2012 [25] | 109,101 | 393 | 193 | 586 | 99.64 | 99.92 | 0.54 |
| Bouaziz et al, 2014 [26] | 109,354 | 140 | 232 | 372 | 99.87 | 99.79 | 0.34 |
| Park et al., 2017 [8] | 109,494 | 79 | 99 | 178 | 99.93 | 99.91 | 0.163 |
| Merah et al., 2018 [27] | 109,316 | 178 | 126 | 304 | 99.84 | 99.88 | 0.28 |

The detectability rate is high and comparable with some methods and outperformed other methods. However, the developed method outperforms all the methods that exist in literature in terms of; (1) simplicity, the proposed method is an easy-to-use method due to its simple framework as compared to other existing methods, and (2) fewer computing resources, the proposed method needs less processing time due to the simple calculation involved. Thus, the developed method represents a reliable and efficient method for real-world application of ECG signals by a wide range of users including medical doctors, scientists, medical labs, etc.

Despite that, the results show a total of 409 detection failures of QRS peaks, which can be classified into five types: (1) failures caused by high-frequency noise as illustrated in Figure 4 and Figure 5 for the record 114 and record 222 respectively which leads to high FP detections of R-peak (2) failures caused by uncertain annotation of true peak in MIT-BIH database that yields to high value of FN in record 116 as illustrated in Figure 6, (3) failures caused by short distance between successive R-peak less than 100 samples for record 116 and 203 as illustrated in Figure 6 and Figure 9, (4) failures caused by large-amplitude artifacts for record 105 which lead to a high number of FN and FP detection as illustrated in Figure 8, and (5) failures caused by reduced amplitude of R-peak that is considerably smaller than the adjacent R-peaks as illustrated in Figure 9. Thus, the detection failures indicate the potential for future work that may address the false detection of R-peak due to short distances between consecutive peaks, high frequencies of noise, large-amplitudes of artifacts, reduced amplitude of R-peak, or uncertain annotation by experts.

**Figure 4.**
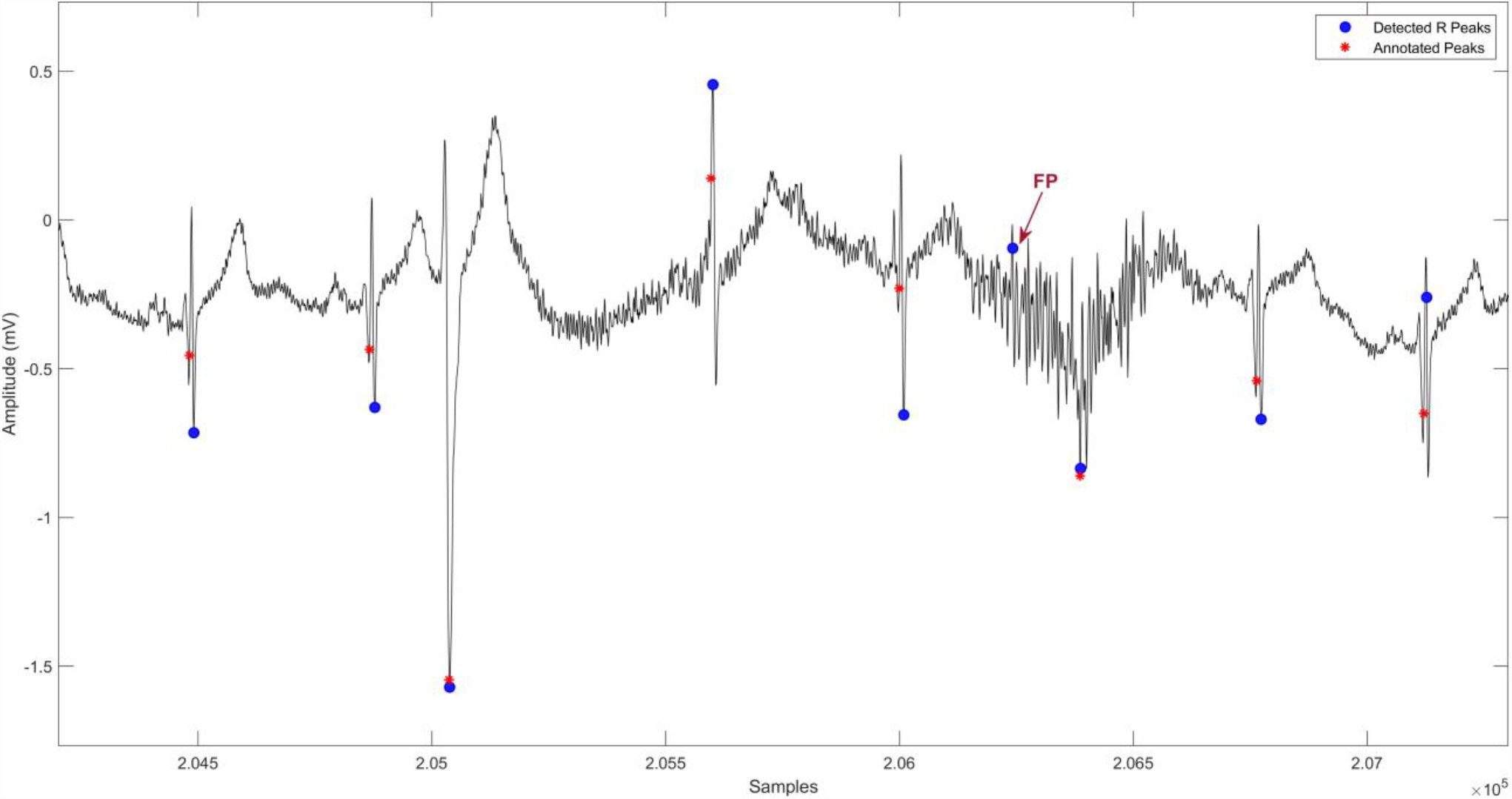
Part of record 114 shows the detection failure (red arrow) of the proposed method caused by high frequency noises

**Figure 5.**
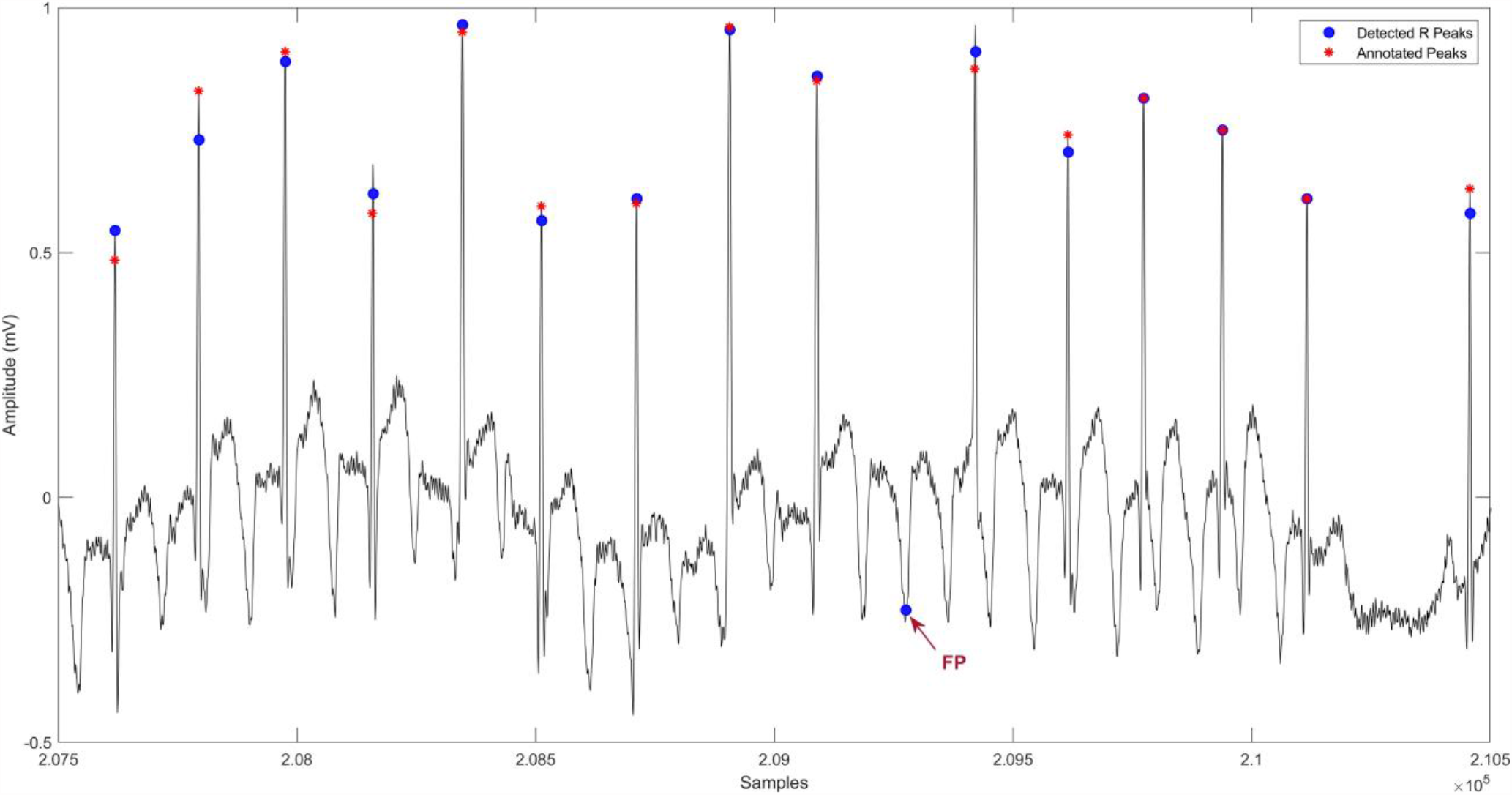
Part of record 222 shows the detection failure (red arrow) of the proposed method caused by high frequency noises and short distances in ECG signal.

**Figure 6.**
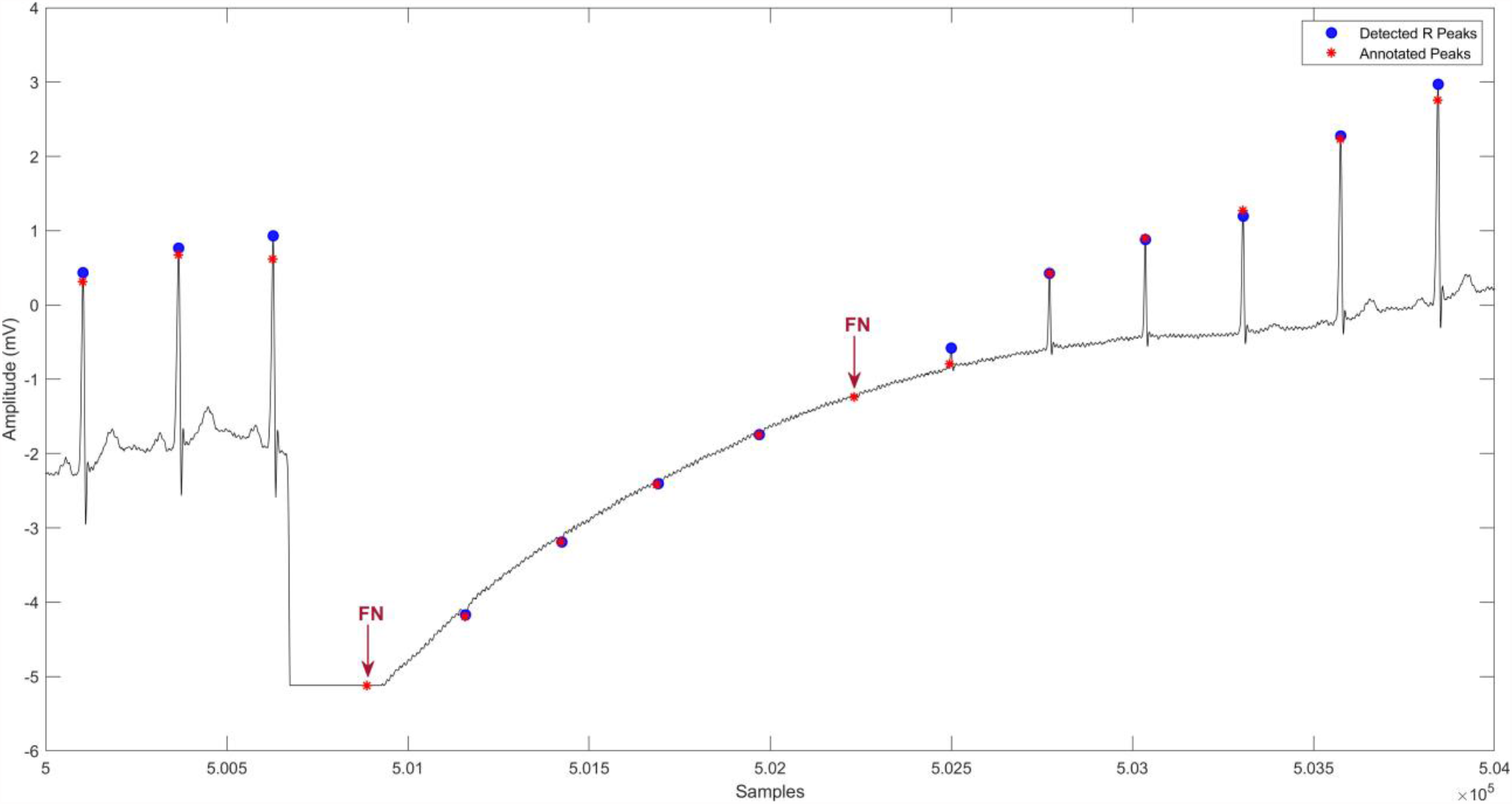
Part of record 116 shows the detection failure of the proposed method caused by Not R-peak being annotated as true peak.

**Figure 7.**
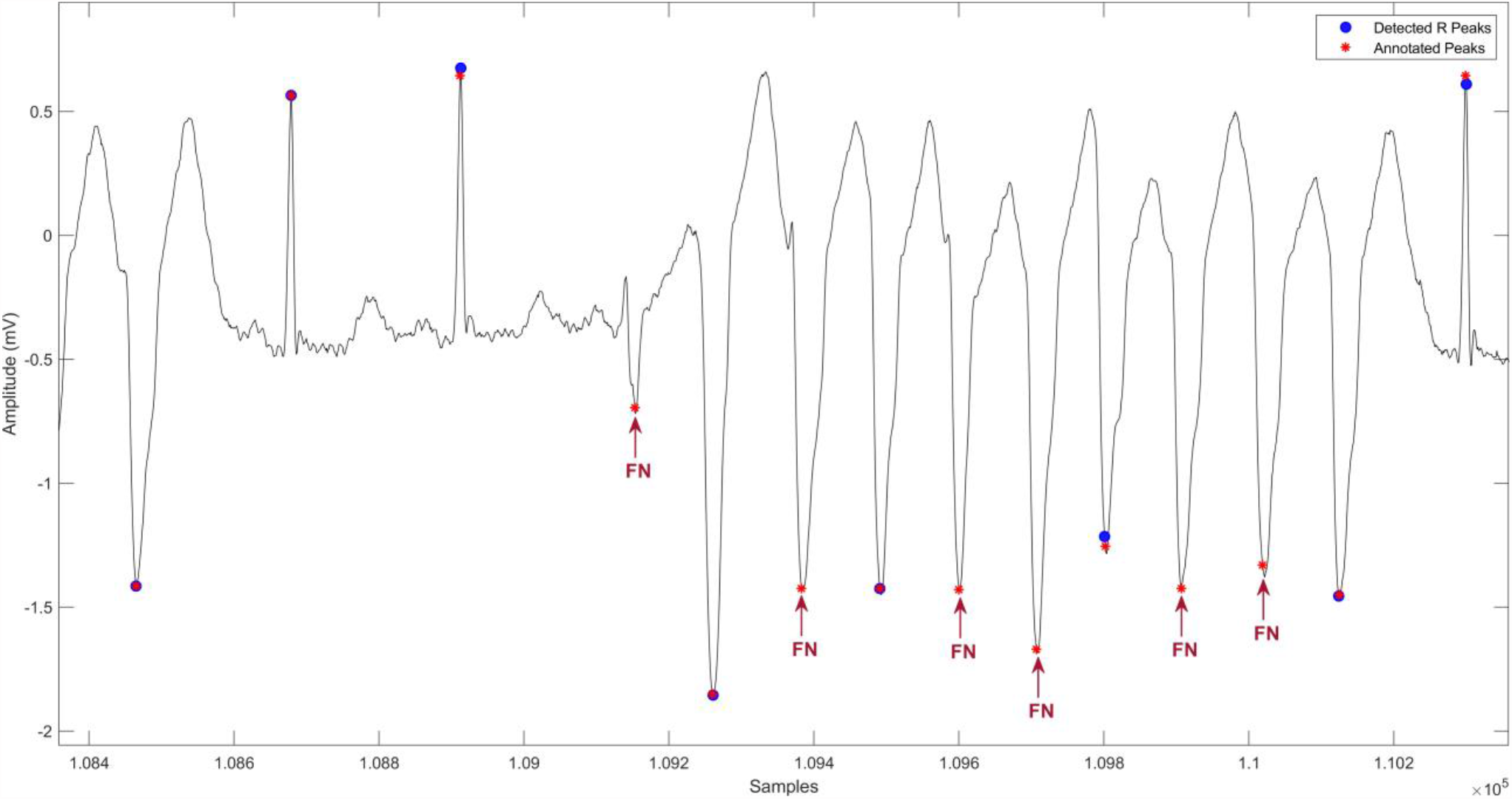
Part of record 205 shows the detection failure of the proposed method caused by the distance shorten then 100 samples between two successive R-peaks.

**Figure 8.**
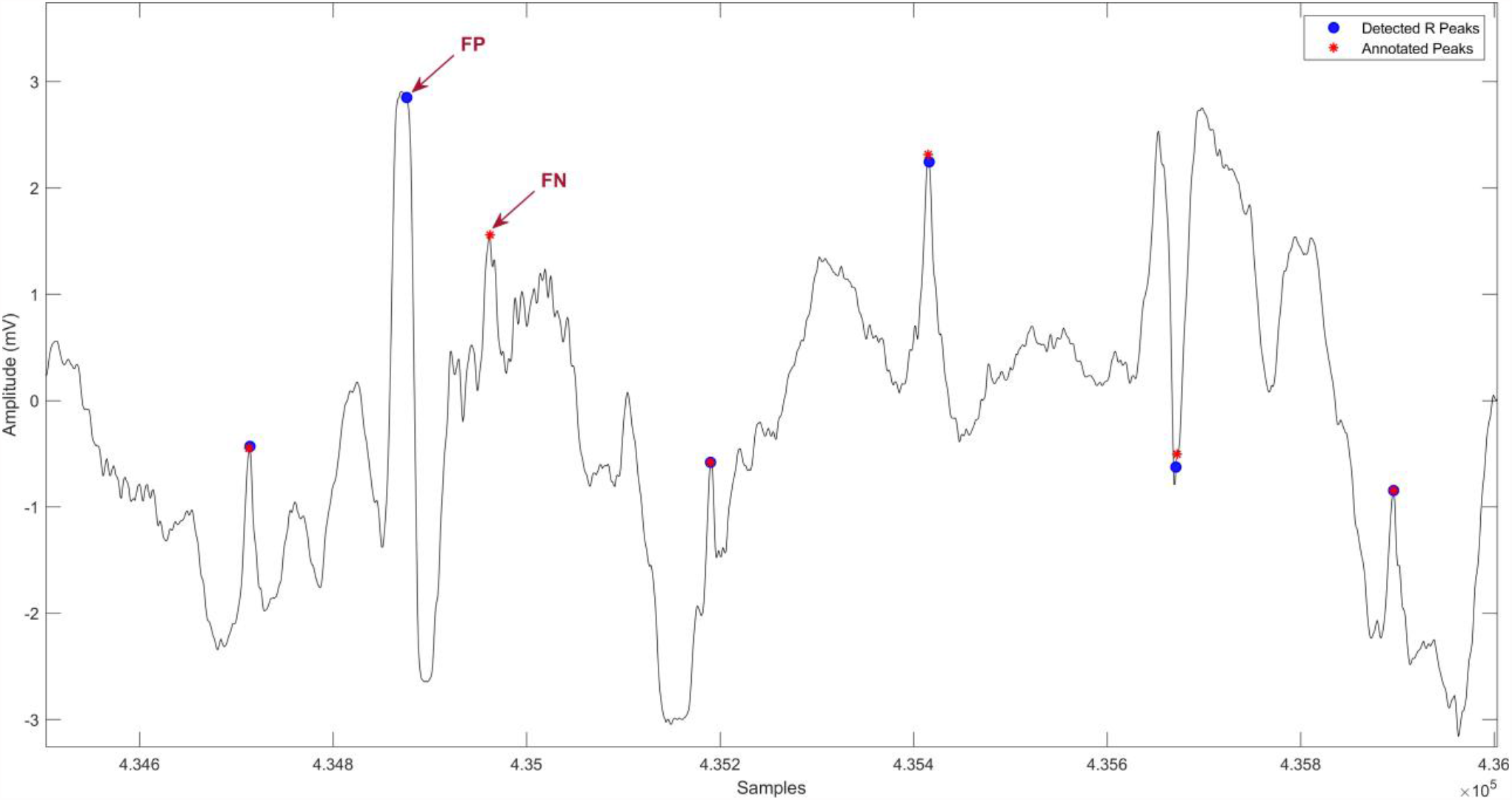
Part of record 105 shows the detection failure of the proposed method caused by artifacts with large amplitude

**Figure 9.**
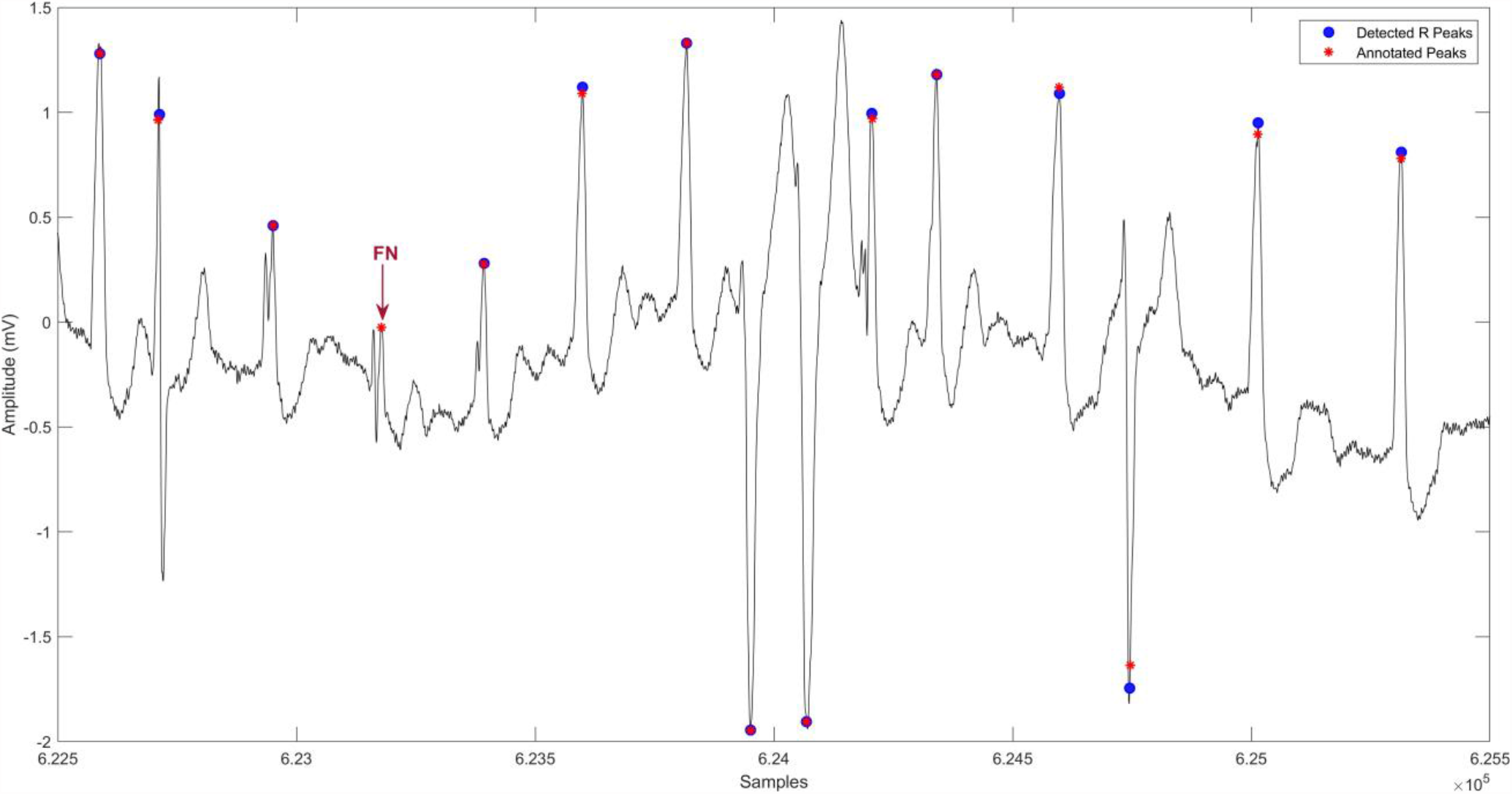
Part of record 203 shows the detection failure of the proposed method caused by the detection P-peak as a R-peak, by short distance between two successive R-peak less 100 samples, and by reduced amplitude of R-peaks.

Performance of proposed filter-less continuous wavelet-based algorithm for R-peak detection in the presence of noise and arrhythmia signals is assessed experimentally using low-quality and noisy signal environments with various levels of signal-to-noise ratio (SNR). The conducted experiment used the noise stress test database (NST) that contains 12 half-hour ECGs and three half-hour noise recordings in the MIT-BIH Noise Stress Test Database (NST) [28]. The ECG signal in NST was created using two clean records from the MIT-BIH Arrhythmia Database (Records 118 and 119) with signal-to-noise ratios of 24, 18, 12, 6, 0, and -6 dB. The NST noise recordings were created with typical ECG recorders/equipment from physically active volunteers. These noise recordings (i.e., Baseline Wander, motion artifacts and EMG) were introduced, after the first 5 minutes of each half-hour ECG recording, in two-minute portions alternating with clean sections.

Table 4 shows the results of analyzing different SNR values that range from 24dB to -6 dB for the same ECG data. These results indicate the performance of the proposed algorithm as compared to Pan-Tompkins algorithm using NST data. In a noisy signal, both algorithms were able to achieve higher sensitivity and predictivity at an SNR range between 24 dB and 18 dB. Even though the detection performance decreased with a higher SNR in the signal, the proposed algorithm outperforms the Pan-Tompkins in identifying the QRS in noisy signal without the use of any filter as shown in Table 4.

**Table 4:** Comparison Performance analysis of the proposed algorithm and Pan-Tompkins Algorithm using <u>the NST</u> database.

| Records | SNR (dB) | Proposed Algorithm |  | Pan-Tompkins Algorithm |  |
| --- | --- | --- | --- | --- | --- |
|  |  | Se (%) | +P (%) | Se (%) | +P (%) |
| 118e24 | 24 (Low Noise) | 100 | 100 | 100 | 99.96 |
| 118e18 | 18 | 100 | 99.43 | 100 | 99.96 |
| 118e12 | 12 | 99.96 | 95.39 | 99.96 | 95.39 |
| 118e06 | 6 | 94.69 | 85.16 | 97.76 | 79.39 |
| 118e00 | 0 | 83.93 | 72.56 | 91.09 | 70.87 |
| 118e_6 | -6 (High Noise) | 76.65 | 66.62 | 71.47 | 60.50 |
| 119e24 | 24 (Low Noise) | 100 | 99.95 | 100 | 100 |
| 119e18 | 18 | 100 | 98.90 | 100 | 99.9 |
| 119e12 | 12 | 100 | 91.52 | 99.95 | 87.92 |
| 119e06 | 6 | 95.42 | 81.16 | 98.09 | 74.28 |
| 119e00 | 0 | 84.85 | 68.93 | 89.63 | 64.34 |
| 119e_6 | -6 (High Noise) | 74.94 | 61.07 | 66.48 | 54.21 |

The results illustrated in Figure 10 highlight the effectiveness of proposed algorithm in detecting QRS under different levels of signal-to-noise ratios from 6 dB to 24 dB with considerably high detection rate. Also, for SNRs lower than 6 dB, the proposed algorithm considerably improves the positive predictivity of QRS by 3 to10 percent as compared to pan-Tompkins. Since the proposed method will not require any filtering, it can be used for monitoring ECG signals for different vendors without the need for any adjustment. The proposed method will also require lower computational resources and thus can be implemented in wearable devices.

**Figure 10.**
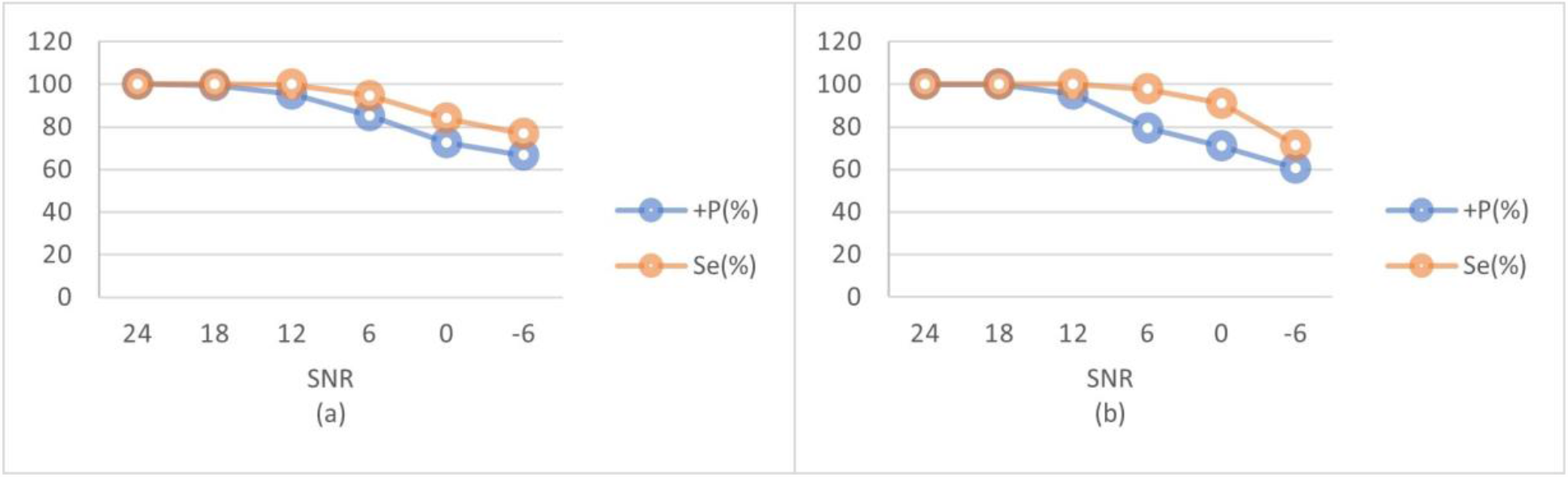
Analysis of the proposed algorithmbehaviour with different SNR. (a) Proposed algorithm, (b) Pan-Tompkins algorithm

## 4. Conclusion

This paper introduces a novel continuous wavelet transform based method for detecting R peaks of an ECG signal. The proposed method utilizes CWT with summation of frequencies to detect the R-peak in ECG signals. The main advantages of the proposed methodology are the simplicity and the fast-processing time of application which allows for rapid ECG signal processing with considerable efficiency and high reliability as compared to existing techniques. The proposed method was applied on several case studies from the MIT-BIH database to validate the performance and to evaluate the accuracy of detection. The results showed the efficiency of the proposed method in analyzing the ECG signals and detecting the R-peaks with considerable detection accuracy with high sensitivity of 99.87%, positive predictability of 99.76% and negligible detection error rate of 0.37%. The comparison of the proposed method with existing techniques in literature indicates its effectiveness and reliability in detecting the R-peaks in ECG signal. In addition to that, the simplicity and minimum computing resources requirement make the developed methods a reliable and efficient R-peak detection method for real world application of ECG signals for a wide range of end-users. The future work may focus on addressing the misidentified R-peaks in terms of small distance between consecutive peaks, high frequencies of noise, and large-amplitude artifacts.

## Data Availability

All data produced are available online at : https://physionet.org/content/ecgiddb/1.0.0/

## References

[1] Cardiovascular diseases (CVDs) (June 2021). URL https://www.who.int/news-room/fact-sheets/detail/cardiovascular-diseases-(cvds)

[2] Y. Sattar, L. Chhabra, Electrocardiogram, StatPearls Publishing, 2021. URL http://www.ncbi.nlm.nih.gov/books/NBK549803/

[3] D. E. Becker, Fundamentals of Electrocardiography Interpretation, Anesth. Prog 53 (2) (2006) 53–64.

[4] I. Vranić, I. Vranić, B. Antić, G. Stojanović, H. Al-Salami, Influence of the Main Filter on QRS-amplitude and Duration in Human Electrocardiogram, Meas. Sci. Rev 19 (1) (2019) 29–34.

[5] S. Nayak, M. Soni, D. Bansal, Filtering techniques for ECG signal processing, in: and others (Ed.), IJREAS. 2., 2012.

[6] Y. Weiting, Z. Runjing, An Improved Self-Adaptive Filter Based on LMS Algorithm for Filtering 50Hz Interference in ECG Signals, 2007 8th International Conference on Electronic Measurement and Instruments (2007) 3–874.

[7] M. Z. Suboh, R. Jaafar, N. A. Nayan, N. H. Harun, Shannon Energy Application for Detection of ECG R-peak using Bandpass Filter and Stockwell Transform Methods, Adv. Electr. Comput. Eng 20 (3) (2020) 41–48.

[8] J. S. Park, S. W. Lee, U. Park, R Peak Detection Method Using Wavelet Transform and Modified Shannon Energy Envelope, J. Healthc. Eng 2017 (2017) 4901017–4901017.

[9] D. Sadhukhan, M. Mitra, Detection of ECG characteristic features using slope thresholding and relative magnitude comparison, 2012 Third International Conference on Emerging Applications of Information Technology (2012) 122–126.

[10] A. Ghaffari, H. Golbayani, M. Ghasemi, A new mathematical based QRS detector using continuous wavelet transform, Comput. Electr. Eng 34 (2) (2008) 81–91.

[11] Ç. P. Dautov, M. S. Özerdem, Wavelet transform and signal denoising using Wavelet method, 2018 26th Signal Processing and Communications Applications Conference (SIU) (2018) 1–4.

[12] R. Golubovski, G. Markoski, E. Golubovska, V. Kokalanov, One implementation of mathematical morphology in medical (ecg) application, Bulletin mathématique de la Société des mathématiciens de la République Macédoine 42 (2) (2018) 101–110. URL http://eprints.ugd.edu.mk/21583/

[13] F. Liu, The Accuracy on the Common Pan-Tompkins Based QRS Detection Methods Through Low-Quality Electrocardiogram Database, J. Med. Imaging Health Inform 7 (2017) 1039–1043.

[14] J. Pan, W. J. Tompkins, A Real-Time QRS Detection Algorithm, IEEE Trans. Biomed. Eng (3) (1985) 230–236.

[15] J. P. Martinez, R. Almeida, S. Olmos, A. P. Rocha, P. Laguna, A wavelet-based ECG delineator: evaluation on standard databases, IEEE Trans. Biomed. Eng 51 (4) (2004) 570–581.

[16] R.-J. Ricardo, E. Martinez-Garcia, T.-C. R. B. Jiri, M.-P. Jolanta, Adaptive Threshold, Wavelet and Hilbert Transform for QRS Detection in Electrocardiogram Signals., in: and others (Ed.), International Conference on P2P, Parallel, Grid, Cloud and Internet Computing, 2017. URL https://www.researchgate.net/publication/319256256_Adaptive_Threshold_Wavelet_and_Hilbert_Transform_for_QRS_Detection_in_Electrocardiogram_Signals

[17] M. S. Manikandan, K. P. Soman, A novel method for detecting R-peaks in electrocardiogram (ECG) signal, Biomed. Signal Process. Control 7 (2) (2012) 118–128.

[18] M. Rahman, M. Milu, A. Anjum, et al., A statistical designing approach to MATLAB based functions for the ECG signal preprocessing, Iran J Comput Sci 2 (2019) 167–178. doi:https://doi.org/10.1007/s42044-019-00035-0.

[19] R. He, K. Wang, Q. Li, et al., A novel method for the detection of R-peaks in ECG based on K-Nearest Neighbors and Particle Swarm Optimization, EURASIP J. Adv. Signal Process 2017 (2017). doi:https://doi.org/10.1186/s13634-017-0519-3.

[20] MIT-BIH Arrhythmia Database (1992). URL https://physionet.org/about/database/

[21] F. Canento, A. Lourenço, H. Silva, A. Fred, et al., Review and Comparison of Real Time Electrocardiogram Segmentation Algorithms for Biometric Applications.

[22] C. C. Lin, H. Y. Chang, Y. H. Huang, C. Y. Yeh, A Novel Wavelet-Based Algorithm for Detection of QRS Complex, Appl. Sci 9 (10) (2019).

[23] M. Yochu, C. Renaud, S. Jacquir, Automatic detection of P, QRS and T patterns in 12 leads ECG signal based on CWT, Biomedical Signal Processing and Control 25 (2016) 46–52. doi:https://doi.org/10.1016/j.bspc.2015.10.011.

[24] A. Chen, Y. Zhang, M. Zhang, W. Liu, S. Chang, H. Wang, H. Wang, J. He, Q. Huang, A Real Time QRS Detection Algorithm Based on ET and PD Controlled Threshold Strategy., Sensors (Basel) 20 (14) (2020). doi:10.3390/s20144003.

[25] Z. Zidelmal, A. Amirou, M. Adnane, A. Belouchrani, QRS detection based on wavelet coefficients, Comput. Methods Programs Biomed 107 (2012) 490–496.

[26] B. Fatiha, D. Boutana, M. Benidir, Multiresolution wavelet-based QRS complex detection algoritm suited toseveral abnormal morphologies, Signal Process. IET 8 (2014) 774–782.

[27] M. Merah, B. H. Abdelmalik, Larbi (2021). [link]. URL https://pubmed.ncbi.nlm.nih.gov/26105724/(accessedNov.17

[28] G. B. Moody, W. E. Muldrow, R. G. Mark, A noise stress test for arrhythmia detectors, Comput. Cardiol., p 11 (1984) 381–384.

